# Development of an Intelligent Predictive Indicator System for Weaning and Extubation Timing in Mechanically Ventilated Patients Based on the Delphi Method

**DOI:** 10.64898/2026.08.02.26359537

**Authors:** Peiyao Li, Haiyan Zhang, Yue Wang, Yang Zhang, Jinjuan Zhang, Xiaowei Meng

## Abstract

**Objective:** To develop an intelligent predictive indicator system for determining weaning and extubation timing in mechanically ventilated patients, providing a theoretical foundation for clinical decision support systems and intelligent assessment in critical care nursing.

**Methods:** A literature review was conducted to identify and extract factors influencing weaning and extubation timing in mechanically ventilated patients, forming the initial item pool for the indicator system. A Delphi expert consultation questionnaire was designed and administered to 21 experts from relevant fields over two rounds. Item screening and revision were performed using the boundary value method, based on the arithmetic mean of importance scores, coefficient of variation (CV), and qualitative expert feedback.

**Results:** The effective response rate was 100% for both rounds. The expert authority coefficient (Cr) was 0.89 in the first round and 0.93 in the second round. Kendall’s W was 0.244 (χ^2^ = 1038.944, P < 0.001) in the first round and 0.079 (χ^2^ = 350.343, P < 0.001) in the second round. Following the first round of evaluation of 203 items—which resulted in 13 additions, 23 modifications, and the separation of weaning/extubation dimensions—the second round comprised 211 evaluation items. The final system encompassed 6 primary indicators and 58 secondary indicators (with tertiary indicators under each), covering the complete clinical trajectory: medical history, ICU admission assessment, pre-weaning/extubation assessment, weaning assessment, extubation assessment, and post-extubation outcomes.

**Conclusion:** This indicator system was developed through a rigorous process with high expert authority and good consensus. The indicators are clinically comprehensive, nursing-operable, and suitable for intelligent modeling. This system can serve as a core framework for risk warning, timing judgment, and intelligent prediction system development for weaning and extubation in mechanically ventilated patients.

## 1. Introduction

Mechanical ventilation is a core life-support intervention in intensive care units (ICUs) for patients with respiratory failure. Weaning and extubation represent critical junctures in the management of mechanically ventilated patients, and precise timing directly impacts treatment outcomes, length of stay, and prognosis [1–2]. In clinical practice, premature extubation may lead to failure, increasing reintubation rates, ventilator-associated pneumonia, and mortality risk. Conversely, delayed extubation prolongs mechanical ventilation duration, elevating the risk of airway injury, diaphragmatic dysfunction, and other complications, while also increasing healthcare resource consumption [3–4]. Currently, clinical assessment of weaning and extubation timing largely depends on physician experience, lacking a standardized and unified evaluation indicator system. Existing indicators are often confined to single dimensions, making comprehensive, dynamic, and precise assessment difficult [5]. Furthermore, critical care nursing lacks standardized support tools for auxiliary assessment of weaning and extubation timing, compromising the targeting and effectiveness of nursing interventions.

With advances in artificial intelligence applications in critical care medicine and nursing, developing intelligent prediction systems for weaning and extubation timing has emerged as a promising solution, with a scientific and systematic indicator system serving as the core foundation for such systems [6–7]. The Delphi method is a qualitative-quantitative research approach that achieves expert consensus through iterative expert consultation and statistical analysis, and has been widely applied in the development of medical indicator systems and scales [8–9]. This study employed the Delphi method, combined with literature review and clinical nursing practice, to construct an intelligent predictive indicator system for weaning and extubation timing in mechanically ventilated patients. The system aims to provide a theoretical basis for precise clinical assessment and intelligent prediction system development, while offering a reference for standardized critical care nursing evaluation.

## 2. Materials and Methods

### 2.1 Establishment of the Research Team

This study was approved by the hospital ethics committee (Approval No. 2025PHB430-001). This study was approved by the hospital ethics committee (Approval No. 2025PHB430-001). All Delphi expert consultation procedures strictly complied with the principles of the Declaration of Helsinki. All invited experts received written research introductions before participation, clearly informed of the research purpose, questionnaire content, anonymous data processing rules and no commercial conflicts of interest. All experts voluntarily participated in the two rounds of consultation and provided implied informed consent by returning the completed questionnaires. All expert personal information was anonymized during data sorting and statistical analysis to fully protect expert privacy.An interdisciplinary research team of 8 members was established, comprising 2 chief physicians from the Department of Critical Care Medicine, 2 associate chief nursing officers, 2 respiratory therapists, 1 master’s-prepared nurse, and 1 master’s-prepared public health specialist. The team’s core responsibilities included: (1) systematic literature search and extraction of factors influencing weaning and extubation timing; (2) clinical expert interviews to supplement and refine indicators based on critical care nursing practice; (3) design of the Delphi expert consultation questionnaire; (4) selection of consultation experts, and standardized distribution, collection, and organization of questionnaires; and (5) statistical analysis of consultation data and indicator system revision based on expert feedback.

### 2.2 Formation of the Initial Indicator Item Pool

A literature search was conducted across CNKI, Wanfang, VIP, PubMed, Embase, and Cochrane Library databases from inception to December 2025. Search terms included: mechanical ventilation, weaning, extubation, timing, prediction, influencing factors, evaluation index, critical care nursing. A total of 1,869 relevant articles were identified. Following independent screening and extraction by two researchers—excluding duplicates, irrelevant articles, and indicators with low clinical practicality—217 items were preliminarily extracted.

Subsequently, 5 senior experts in critical care medicine, critical care nursing, and nursing management were interviewed using a semi-structured approach. Indicators were supplemented, merged, and revised based on clinical practice and assessment needs—for example, merging duplicate respiratory function indicators, adding nursing-relevant symptom assessment indicators, and removing indicators difficult to implement in primary care settings. The final initial item pool comprised 5 primary indicators, 46 secondary indicators, and 153 tertiary indicators (204 items total). The primary indicators included: medical history (past and present), ICU admission assessment, pre-weaning/extubation assessment, weaning/extubation assessment, and post-weaning/extubation outcomes.

### 2.3 Design of the Expert Consultation Questionnaire

The questionnaire was designed following Delphi method guidelines [9–10], incorporating four sections. The overall content was concise and logically structured for ease of completion:

Section 1 Introduction: This section presented the background, objectives, significance, indicator system framework, and instructions for completing the questionnaire. It clarified the rating criteria and the format for providing recommendations, and emphasized the anonymity of responses and the professionalism of the results.

Section 2 Questionnaire body: The initial indicator item pool was presented, with each item rated on a 5-point Likert scale for importance (1 = not important to 5 = very important). A comment section was provided for each item, allowing experts to propose specific recommendations regarding wording, classification, and addition or deletion of items based on clinical and nursing considerations.

Section 3 Expert background: This section collected information on gender, age, education, professional title, specialty area, years of experience in critical care medicine/nursing, and prior involvement in weaning/extubation-related guideline development, clinical research, or care pathway construction.

Section 4 Expert self-assessment: Familiarity with the subject was rated on a 5-level scale: very familiar (assigned value 1.0), familiar (0.8), generally familiar (0.6), not very familiar (0.4), and unfamiliar (0.2). Judgment basis was categorized into four types: practical experience (large = 0.5, medium = 0.4, small = 0.3), theoretical analysis (large = 0.3, medium = 0.2, small = 0.1), literature reference (large = 0.1, medium = 0.1, small = 0.1), and subjective judgment (large = 0.1, medium = 0.1, small = 0.1). These were used to calculate the judgment basis coefficient (Ca) and familiarity coefficient (Cs).

### 2.4 Expert Selection

Selection criteria, based on Delphi method principles of representativeness, authority, engagement, and nursing relevance [9], were established in accordance with the study theme and critical care nursing research context: (1) ≥10 years of experience in critical care medicine, respiratory therapy, critical care nursing, or nursing management; (2) intermediate or higher professional title (senior preferred), with ≥50% from critical care nursing to ensure alignment with clinical nursing practice; (3) familiarity with clinical management and nursing assessment of weaning and extubation in mechanically ventilated patients; (4) prior involvement in relevant research, guideline development, clinical pathway construction, or nursing quality indicator development (preferred); and (5) voluntary participation with commitment to complete both consultation rounds and provide specific, substantive recommendations.

Based on these criteria, 21 experts from tertiary hospitals across multiple provinces were selected, covering critical care medicine, respiratory medicine, respiratory therapy, critical care nursing, and nursing management, with 12 from critical care nursing-related fields to ensure alignment with clinical nursing practice.

### 2.5 Delphi Consultation Process

Questionnaires were distributed and collected via WeChat and email. The first round was conducted in February 2026. Along with questionnaire distribution, one-on-one communication was maintained to clarify completion requirements, rating criteria, and the 1-week response deadline, with timely responses to expert queries.

Following the first round, the research team performed standardized statistical analyses, calculating importance scores and CVs for each indicator. Expert recommendations were systematically reviewed. Revisions were made based on scientific validity, clinical practicality, and nursing operability: (1) retaining indicators with mean importance ≥4.00 and CV ≤0.25; (2) revising indicators with unclear wording or classification; (3) incorporating highly endorsed supplementary clinical and nursing indicators; and (4) retaining low-scoring (<4.00) or high-disagreement (CV > 0.25) indicators for re-evaluation in the second round.

Based on first-round results, the revised second-round questionnaire was distributed to the same expert panel in April 2026. Following collection, statistical analysis and qualitative review were repeated. After two rounds, expert consensus was achieved, and the consultation was terminated.

### 2.6 Statistical Analysis

Data were analyzed using Excel 2019 and SPSS 26.0. Statistical methods conformed to Delphi method standards [8–11]:

#### Expert engagement

Effective response rate (effective responses / distributed questionnaires×100%) and recommendation rate (experts providing recommendations / effective responses × 100%).

#### Expert authority

Authority coefficient Cr= (Ca+Cs)/2, with Cr ≥ 0.70 considered acceptable.

#### Expert consensus

Kendall’s W coefficient for overall agreement; CV for single-item agreement. The χ^2^ test was performed for W, with P < 0.05 considered statistically significant.

#### Item screening

Boundary value method (mean – SD), with items scoring below the boundary deleted, and items near the boundary retained or deleted based on expert feedback and clinical relevance.

## 3. Results

### 3.1 Expert Demographics

All 21 experts were from tertiary hospitals, covering critical care medicine, critical care nursing, and related fields. Demographic characteristics are summarized in Table 1.

**Table 1.** Demographic Characteristics of Expert Panel (n = 21)

| Characteristic | Category | n | Percentage (%) |
| --- | --- | --- | --- |
| Age (years) | 30–39 | 6 | 28.57 |
|  | 40–49 | 10 | 47.62 |
|  | 50–59 | 5 | 23.81 |
| Years of Experience | 10–19 | 9 | 42.86 |
|  | 20–29 | 5 | 23.81 |
|  | 30–39 | 7 | 33.33 |
| Professional Title | Chief Physician | 1 | 4.76 |
|  | Chief Nursing Officer | 6 | 28.57 |
|  | Charge Nurse | 2 | 9.52 |
|  | Charge Technologist | 1 | 4.76 |
|  | Associate Chief Nursing Officer | 8 | 38.10 |
|  | Associate Professor | 3 | 14.29 |
| Specialty Area | Critical Care | 10 | 47.62 |
|  | Critical Care & Respiratory | 3 | 14.29 |
|  | Respiratory Therapy | 1 | 4.76 |
|  | Nursing Management | 1 | 4.76 |
|  | Critical Care & Nursing Management | 6 | 28.57 |
| Education | Associate Degree | 1 | 4.76 |
|  | Bachelor's | 13 | 61.90 |
|  | Master's | 5 | 23.81 |
|  | Doctoral or above | 2 | 9.52 |

### 3.2 Delphi Consultation Evaluation

#### 3.2.1 Expert Engagement

Both rounds achieved a 100% effective response rate (21/21). In the first round, 15 experts (71.43%) provided specific recommendations; in the second round, 18 experts (85.71%) provided recommendations, with second-round recommendations more closely aligned with clinical nursing practice.

#### 3.2.2 Expert Authority

The authority coefficients, judgment basis coefficients, and familiarity coefficients for both rounds all met the acceptable evaluation standard of Cr ≥ 0.70. The familiarity coefficient in the second round showed a notable improvement compared to the first round, while the judgment basis coefficient remained stable. This indicates that, based on the feedback from the first round and the indicator system revision process, experts developed a deeper understanding of the indicator construction logic and critical care nursing requirements, with corresponding improvements in the accuracy and consistency of evaluation results (Table 2).

**Table 2.** Expert Authority Coefficients Across Two Rounds.

| Round | Judgment Basis (Ca) | Familiarity (Cs) | Authority Coefficient (Cr) |
| --- | --- | --- | --- |
| 1 | 0.98 | 0.80 | 0.89 |
| 2 | 0.91 | 0.96 | 0.93 |

#### 3.2.3 Expert Consensus

Kendall’s W and χ^2^ tests for both rounds showed P < 0.001, indicating that expert evaluations of indicator importance demonstrated statistically significant consistency, with overall consensus at an acceptable level. The decrease in W from Round 1 to Round 2 is primarily attributable to the addition of 13 clinical and nursing indicators and the reclassification of some indicators following the first round, increasing the total item count from 203 to 211. The newly added indicators and reclassified structure resulted in moderate differences in expert opinion, a phenomenon consistent with the methodological characteristics of multi-round Delphi studies [10–11], rather than indicating evaluation bias (Table 3).

**Table 3.** Expert Consensus Across Two Rounds.

| Round | n | Items Evaluated | W | $\chi^2$ | df | P |
| --- | --- | --- | --- | --- | --- | --- |
| 1 | 21 | 203 | 0.244 | 1038.944 | 203 | <0.001 |
| 2 | 21 | 211 | 0.079 | 350.343 | 211 | <0.001 |

At the item level, in the first round, 10 core indicators—including COPD, myasthenia gravis, respiratory failure, and difficult airway—achieved CV = 0.00, indicating complete agreement among experts. Indicators such as sex (CV = 0.33), blood glucose (CV = 0.30), and blood urea nitrogen (CV = 0.25) showed CV > 0.25, indicating substantial disagreement. In the second round, most indicators had CV < 0.25; newly added nursing-related indicators—including pain score, RASS score, and cuff leak test—were highly endorsed, all with CV < 0.15. Sex (CV = 0.24), blood glucose (CV = 0.30), and blood urea nitrogen (CV = 0.25) remained somewhat discordant but showed decreasing trends compared to the first round, indicating gradual convergence of expert opinions.

### 3.3 Indicator System Revision and Finalization

#### 3.3.1 Round 1 Revisions

Based on first-round statistical results and expert recommendations, combined with clinical nursing practice, the initial indicator system was systematically revised:

Retained: 186 indicators with mean importance ≥4.00 and CV ≤ 0.25—such as COPD, rapid shallow breathing index (RSBI), weaning failure, and extubation failure—all of which are key indicators in clinical diagnosis, treatment, and nursing assessment.

Revised: 23 indicators with unclear wording or classification—for example, “invasive blood pressure” was revised to “blood pressure (including invasive/noninvasive),” “albumin” was revised to “serum albumin (ALB),” and “ventilator mode” was adjusted to “ventilator parameters”—to align indicator terminology more closely with clinical nursing practice.

Added: 13 indicators highly endorsed by experts, spanning respiratory function, inflammatory/metabolic, safety, and nursing assessment dimensions—including bedside driving pressure measurement before extubation, neutrophil-to-lymphocyte ratio (NLR), cuff leak test, RASS score, and pain score.

Carried forward for re-evaluation: 6 low-scoring, high-disagreement indicators (sex, blood glucose, etc.) were retained for re-evaluation in the second round.

Reclassified: Disease-related indicators were regrouped by neurological, respiratory, and circulatory systems; creatinine and blood urea nitrogen from blood gas parameters were reassigned to laboratory indicators to avoid classification confusion that could lead to nursing assessment bias.

#### 3.3.2 Round 2 Finalization

Following the second round of expert consultation, the revised indicator system received increased endorsement; newly added nursing indicators all achieved mean importance scores ≥4.60. Only a small number of indicators—sex, blood glucose, and blood urea nitrogen—remained somewhat discordant. The research team completed the final optimization of the indicator system based on the combined recommendations from both rounds, clinical critical care scenarios, and intelligent prediction system development requirements:

The original primary indicator “Weaning/Extubation Assessment” was split into “Weaning Assessment” and “Extubation Assessment,” with the addition of secondary indicators related to safety assurance and weaning tolerance assessment.Items with low clinical practicality were removed; psychological and swallowing function assessment indicators were added to align with humanistic critical care and clinical assessment needs.All indicator terminology was standardized, with clear clinical definitions, nursing assessment methods, and judgment criteria to enhance clinical operability.

The final indicator system comprises 6 primary indicators and 58 secondary indicators (with tertiary indicators under each). The primary indicators are: (1) Medical History, (2) ICU Admission Assessment, (3) Pre-Weaning/Extubation Assessment, (4) Weaning Assessment, (5) Extubation Assessment, and (6) Post-Weaning/Extubation Outcomes. Core indicators—including difficult airway, RSBI, maximum inspiratory pressure (MIP), weaning failure, extubation failure, and cuff leak test—achieved full or near-full scores with CV < 0.10, reflecting extremely high expert consensus, and all are key indicators in critical care nursing clinical assessment.

## 4. Discussion

### 4.1 Reliability of the Delphi Consultation

In this study, the 100% effective response rates in both rounds, along with the increase in recommendation rate from 71.43% in the first round to 85.71% in the second round, indicate high levels of expert engagement. The expert authority coefficients (Cr = 0.89 in the first round and 0.93 in the second round) significantly exceeded the 0.70 reliability threshold; notably, the familiarity coefficient increased from 0.80 to 0.96, while the judgment basis coefficient remained consistently above 0.90, indicating that experts possessed sufficient judgment basis and that their familiarity with the subject matter increased substantially over the course of the consultation process. The 21 experts were recruited from tertiary hospitals across China, with specialties covering critical care medicine, respiratory therapy, critical care nursing, and nursing management. This multidisciplinary composition effectively mitigated single-perspective bias and ensured the authority and comprehensiveness of the opinions.

In terms of overall consensus, Kendall’s W χ^2^ tests for both rounds showed P < 0.001, indicating statistically consistent expert evaluations at an acceptable level. The slight decrease in W in the second round is attributable to the addition of 13 clinical and nursing indicators and the reclassification of some indicators following the first round, increasing the total item count from 203 to 211. The newly added indicators and reclassified structure resulted in moderate differences in expert opinion—a phenomenon consistent with the methodological characteristics of multi-round Delphi studies [10–11], rather than indicating evaluation bias. Core indicators—including COPD, difficult airway, RSBI, and cuff leak test—achieved CV = 0.00, reflecting complete expert consensus. Indicators such as sex, blood glucose, and BUN showed decreasing divergence (sex: CV 0.33 → 0.24); the remaining disagreement does not compromise the overall consensus foundation, primarily stemming from insufficient evidence for direct causality and differences in assessment perspectives between specialty areas.

### 4.2 Characteristics and Clinical Nursing Value of the Indicator System

This indicator system has four distinguishing features. First, it provides comprehensive, whole-process coverage, spanning from admission to post-extubation outcomes, integrating respiratory mechanics, vital signs, blood gases, laboratory tests, imaging, psychological, and nursing assessment indicators. Second, it has strong clinical operability—items with low clinical practicality were removed following expert consultation, and urgently needed bedside indicators—including bedside driving pressure, SBT, and cuff leak test—were added, with clear assessment methods and judgment cutoffs defined for each indicator to facilitate bedside application. Third, it has explicit nursing orientation—the system incorporates nursing-sensitive indicators including RASS score, pain assessment, consciousness evaluation, swallowing function, and psychological status assessment; the cuff leak test is designated as a safety assessment item, making the system simultaneously applicable to both clinical decision-making and nursing assessment needs, and providing a concrete tool for standardized critical care nursing evaluation. Fourth, it has intelligent system compatibility—the “Weaning/Extubation Assessment” dimension was split into separate “Weaning Assessment” and “Extubation Assessment” dimensions, with all indicators standardized and structurally named with clear time points, meeting the data format requirements of machine learning algorithms. Core indicators that received full scores from experts—COPD, RSBI, MIP, etc.—constitute the core assessment targets, providing clear direction for rapid clinical decision-making and subsequent feature selection in prediction models.

### 4.3 Analysis of Discordant Indicators

Sex, blood glucose, and blood urea nitrogen remained somewhat discordant after two rounds of consultation (sex: CV 0.33 → 0.24, mean score 3.67 → 4.14), primarily reflecting two factors. First, there is insufficient evidence for a direct causal relationship between sex and weaning/extubation outcomes; most experts considered that sex exerts its influence indirectly through physical condition and underlying comorbidities, rather than as an independent influencing factor. Second, physicians and nursing experts have different assessment priorities—physicians tend to focus on the impact of blood glucose and renal function on overall prognosis, while nursing experts emphasize their reference value for immediate weaning/extubation outcomes, and these indicators are susceptible to interference from clinical interventions. Accordingly, these indicators are provisionally designated as “pending verification indicators” in this study, and future large-sample empirical studies are recommended to clarify their supplementary predictive value and establish reference thresholds.

### 4.4 Limitations and Future Directions

This study has several limitations. First, all consultation experts were recruited from tertiary hospitals; perspectives from primary care settings were not included, and the generalizability of the system to lower-resource settings requires validation. Second, indicator weights have not yet been assigned—future application of the Analytic Hierarchy Process (AHP) is planned, particularly for weighting nursing-sensitive indicators. Third, clinical validation of predictive performance and nursing operability has not yet been conducted. Future plans include a multicenter prospective validation study with a large sample size, development of an intelligent prediction system with integrated nursing assessment modules, and establishment of standardized nursing assessment pathways for weaning and extubation to evaluate the system’s impact on nursing quality and patient outcomes.

## 5. Conclusion

This study successfully developed an intelligent predictive indicator system for weaning and extubation timing in mechanically ventilated patients, comprising 6 primary indicators, 58 secondary indicators, and tertiary indicators under each. By incorporating nursing-sensitive indicators—RASS score, pain assessment, swallowing function, and psychological factors—the system provides quantifiable assessment tools for critical care nursing in weaning and extubation decision-making, thereby enhancing the standardization of nursing assessment. Core predictors including COPD, difficult airway, RSBI, and MIP were identified with complete expert consensus, and all indicators were standardized and structurally named to meet the data requirements of intelligent prediction systems. The system achieves a balance of scientific rigor, nursing practicality, and intelligent adaptability, providing an actionable decision-making framework for improving weaning and extubation success rates and promoting standardized assessment and intelligent transformation in critical care nursing. Following weight assignment and clinical validation, the system’s practical value and predictive performance will be further established.

## Data Availability

N/A

## Ethics Approval

Ethics Committee of Peking University People’s Hospital (Approval No.: 2025PHB430-001)

## Date of Completion

April 2026

